# Characteristics and outcomes of patients treated with sotrovimab to prevent progression to severe COVID-19 in Belgium

**DOI:** 10.1101/2023.12.14.23298578

**Authors:** Myriam Drysdale, Thor Hautekiet, Moushmi Singh, Joris Hautekiet, Linda Ludikhuyze, Vishal Patel, Daniel C Gibbons, Dorothée De Roeck, Kirsten Colpaert, Emily J Lloyd, Eva Van Braeckel

**Affiliations:** GSK, Brentford, UK; Department of Respiratory Medicine, Ghent University Hospital, Ghent, Belgium; IQVIA Belux, Zaventem, Belgium; GSK, Wavre, Belgium; Department of Intensive Care, Ghent University Hospital, Ghent, Belgium; Respiratory Infection and Defense Lab, Department of Internal Medicine and Pediatrics, Faculty of Medicine and Health Sciences, Ghent University, Ghent, Belgium

**Author notes:** Correspondence: Myriam Drysdale. Myriam Drysdale and Thor Hautekiet contributed equally to the manuscript. Affiliation correct at time of study (Joris Hautekiet, Vishal Patel).

**Keywords:** COVID-19, monoclonal antibody, Omicron BA.1, Omicron BA.2, SARS-CoV-2, sotrovimab

## Abstract

**Background:** Sotrovimab, a dual-action, engineered human monoclonal antibody, has been demonstrated to significantly reduce the risk of hospitalization and death in high-risk patients with COVID-19. Here, we describe the real-world use of, and outcomes from, sotrovimab treatment in Belgium during the Delta and Omicron waves among patients with COVID-19 at high risk of developing severe disease.

**Methods:** This was a multicentric, single-arm observational cohort study of non-hospitalized patients receiving outpatient sotrovimab treatment between 1 November 2021 and 2 August 2022. We performed a retrospective analysis of hospital, pharmacy and administrative data from nine hospitals in Belgium. The primary outcomes were all-cause and COVID-19-related hospitalizations and all-cause deaths during the 29-day acute follow-up period from first administration of sotrovimab.

**Results:** A total of 634 patients were included in the analysis (63.4% aged <65 years; 50.3% male). A high proportion (67.7%; *n* = 429/634) of patients were immunocompromised, with 36.9% (*n* = 234/634) actively treated for malignancy. During the 29-day acute period, 12.5% (*n* = 79/634) of sotrovimab-treated patients were hospitalized due to any cause (median duration 4 days; median time to hospitalization 14 days) and 1.1% (*n* = 7/634) died due to any cause. In total, 0.8% (*n* = 5/634) of patients were admitted to an intensive care unit (ICU). COVID-19-related hospitalization was experienced by 2.5% (*n* = 16/634) of patients (median duration 10 days; median time to hospitalization 10.5 days), with 0.5% (*n* = 3/634) of patients admitted to an ICU. COVID-19-related hospitalization was experienced by 6.3% (*n* = 3/48) of patients during Delta predominance (04/11/2021–23/12/2021), 6.3% (*n* = 1/16) of patients during Delta/BA.1 codominance (24/12/2021–01/01/2022), 1.4% (*n* = 3/218) of patients during BA.1 predominance (02/01/2022–09/02/2022), 2.1% (*n* = 2/97) of patients during BA.1/BA.2 codominance (10/02/2022–07/03/2022) and 2.7% (*n* = 7/255) of patients during BA.2/BA.5 codominance (08/03/2022–02/08/2022).

**Conclusions:** This observational study demonstrated consistently low rates of COVID-19-related hospitalizations and all-cause deaths in sotrovimab-treated patients during the Omicron subvariant periods in Belgium, despite over two-thirds of the study population being immunocompromised. Comparative effectiveness studies are warranted to confirm sotrovimab effectiveness in highly immunocompromised patients with COVID-19.

## Background

Coronavirus disease 2019 (COVID-19) is caused by infection with severe acute respiratory syndrome coronavirus 2 (SARS-CoV-2); the rapid global spread of SARS-CoV-2 resulted in the declaration of a pandemic by the World Health Organization in March 2020 [1]. Globally, as of September 2023, there have been over 770 million confirmed cases of COVID-19, including nearly 7 million deaths [2]. In Belgium, almost 158,000 hospitalizations and 34,000 deaths have been attributed to COVID-19 during the same period [2, 3].

Older individuals or those with immunosuppression, cancer, obesity, diabetes, or chronic kidney, lung, liver or cardiovascular disease are known to be at higher risk of developing severe COVID-19, which may result in hospitalization or death [4–6].

Sotrovimab is a dual-action, engineered human immunoglobulin G1κ monoclonal antibody (mAb) derived from the parental mAb S309, a potent neutralising mAb directed against the spike protein of SARS-CoV-2 [7–10]. Sotrovimab (administered as a single 500 mg intravenous dose) was shown in COMET-ICE, a randomized clinical trial, to significantly reduce the risk of all-cause >24-hour hospitalization or death compared with placebo in high-risk patients with COVID-19 [11].

As a result of positive findings from COMET-ICE, sotrovimab received marketing authorization from the European Medicines Agency (EMA) in December 2021 for use in adults and adolescents (aged ≥12 years and weighing ≥40 kg) with COVID-19 who do not require oxygen supplementation and are at increased risk of progressing to severe COVID-19 [12]. In Belgium, use of sotrovimab began prior to EMA approval as a result of a procurement framework contract issued by the European Commission in July 2021 that enabled participating European Union member states, including Belgium, to purchase sotrovimab prior to its marketing authorization [13, 14]. At the time of the study, mAbs (including sotrovimab) were recommended for the treatment of mild-to-moderate COVID-19 among patients in Belgium at high risk of complications, following authorization by a multidisciplinary team including at least one infectious disease physician and one immunologist [15]. Due to limited supplies of mAbs, immunocompromised patients were considered the highest priority for mAb treatment [15].

Sotrovimab has been granted marketing authorization in the European Union, Norway, Iceland [16], Australia (conditional) [17], United Kingdom (conditional) [18], Saudi Arabia (conditional) [19] and Switzerland (conditional) [19]. In Japan, a Special Approval for Emergency designation has been granted [20]. Since the approval of sotrovimab, waves of new SARS-CoV-2 variants of concern have emerged, including the Delta variant and the Omicron BA.1, BA.2 and BA.5 subvariants; the latter became dominant in Belgium in June 2022 [21–23]. *In vitro* neutralization studies have demonstrated a moderate fold shift in activity of sotrovimab against Omicron BA.2 relative to wild-type SARS-CoV-2 (16-fold change in EC_50_ relative to wild-type) [24]. A similar reduction in activity was reported for sotrovimab against BA.5 (22.6-fold change in EC_50_ relative to wild-type) [24]. Consequently, on 28 April 2022, the Federal Agency for Medicines and Health Products regulatory body in Belgium took the decision to no longer recommend the use of sotrovimab [25], although use continued among very-high-risk and/or immunocompromised patients.

In the absence of a clinical trial conducted during the Omicron predominance period, it remains uncertain how *in vitro* antibody neutralization activity correlates with clinical effectiveness, especially for dual-action antibodies like sotrovimab that have potent effector function, including antibody-dependent cellular cytotoxicity and antibody-dependent cellular phagocytosis [10, 26, 27]. Real-world studies are an important source of data with which to assess clinical outcomes in patients treated with sotrovimab within the evolving variant landscape.

Here, we describe the real-world use of, and outcomes from, sotrovimab treatment for the management of patients with COVID-19 at high-risk of developing severe disease between 1 November 2021 and 2 August 2022 in Belgium, when the Delta variant and Omicron BA.1, BA.2 and BA.5 subvariants were predominant.

## Methods

### Study design and data source

In this single-arm observational cohort study, data were collected retrospectively from nine centres (including two university hospitals) located in Flanders, Brussels and Wallonia that administered sotrovimab in an outpatient setting from 1 November 2021 up to and including 2 August 2022. Patient-level data were collected and aggregated from three main sources: 1) registered administrative, medical and nursing data, which are fully structured and standardized across all hospitals and therefore require no specific technology to be implemented for data retrieval; 2) structured and standardized hospital pharmacy or invoicing data using predefined Anatomical Therapeutic Chemical (ATC) codes to capture the date of sotrovimab administration; 3) administrative data regarding hospital admission and transfer.

The index date (Day 1) was defined as the date of first treatment administration with sotrovimab. The baseline period was defined as the 1 year prior to index for assessment of prior comorbidities. All-cause and COVID-19-related outcomes were reported during the acute period, defined as the 29-day follow-up period starting on the index date. End of follow-up was defined as either the end of the acute period, death or the study end (31 August 2022), whichever occurred first.

This study complies with all applicable laws regarding subject privacy. No direct subject contact or primary collection of individual human subject data occurred. Study results were in tabular form and were aggregate analyses that omitted subject identification; therefore, informed consent was not required. Ethics committee approval was obtained from each of the participating centres, with the exception of AZ Maria Middelares, AZ Groeninge and Jan Yperman where approval was provided by a data access committee.

### Study population

Non-hospitalized patients infected with SARS-CoV-2 were eligible for inclusion if they were aged ≥12 years on the index date and received outpatient treatment with sotrovimab for the first time during the study period.

Patients were excluded if they received sotrovimab in an inpatient setting and/or received remdesivir as an early treatment in an outpatient setting during the acute period. However, in three of the study centres, patients routinely stayed in hospital overnight after sotrovimab administration for logistical (i.e. for monitoring post treatment administration) rather than medical reasons; therefore, the exclusion criteria were adapted for these centres to include patients discharged the day after sotrovimab administration.

### Study outcomes

Patient demographics and clinical characteristics were collected based on the presence of selected International Classification of Disease version 10 (ICD-10) and ATC codes (Tables S1–S3) during any hospital visit that occurred in the 12 months prior to the index date.

These included age, sex, and comorbidities and risk factors that may predispose patients to severe COVID-19 outcomes, as follows: immunocompromised status (human immunodeficiency virus infection/acquired immunodeficiency syndrome, treatment for solid tumours, haematological malignancy, solid organ or haematopoietic stem cell transplantation, primary immune deficiency, receiving immunosuppressive drugs [see **Table S1**], sickle-cell anaemia, major thalassaemia, asplenia); other risk factors or comorbidities of interest (age ≥65 years old, obesity, type 1 or type 2 diabetes mellitus, chronic lung disease including asthma and cystic fibrosis, cardiovascular disease including uncontrolled hypertension, chronic kidney disease [CKD; estimated glomerular filtration rate <30 ml/min] including treatment via haemodialysis, chronic liver disease [Child–Pugh B or C], chronic neurological disease). These comorbidities and risk factors for severe COVID-19 aligned with those reported by the Scientific Institute for Public Health of the Federal Belgian State (Sciensano) and the US Centers for Disease Control and Prevention [5, 6].

The clinical outcomes of this study included the number and proportion of all-cause and COVID-19-related hospitalizations and all-cause deaths during the acute period (29 days after the index date). COVID-19-related hospitalizations were defined as having the following ICD-10 codes registered as the verified admission diagnosis: B34.2, B97.2 or U07.1 (**Table S2**). In a *post hoc* analysis, cause of death data were collected for patients at Ghent University Hospital and the proportion of COVID-19-related deaths was determined; only all-cause deaths are reported for the other centres.

The number and proportion of all-cause and COVID-19-related hospitalizations and all-cause deaths among sotrovimab-treated patients were reported for the overall population and stratified by week and month, as well by the period of SARS-CoV-2 variant predominance in Belgium [28], as follows: Delta predominance period, 4 November 2021– 23 December 2021; Delta/BA.1 codominance period, 24 December 2021–1 January 2022; BA.1 predominance period, 2 January 2022–9 February 2022; BA.1/BA.2 codominance period, 10 February 2022–7 March 2022; BA.2/BA.5 codominance period, 8 March 2022–2 August 2022 (Fig. 1). The study design did not enable collection of polymerase chain reaction results in order to identify the SARS-CoV-2 variant strains that patients were infected with. Rather, to map the 29-day outcomes with the circulating variants, we used a proxy based on the most prevalent strains circulating in Belgium per week, as provided by Sciensano [28].

**Fig. 1.**
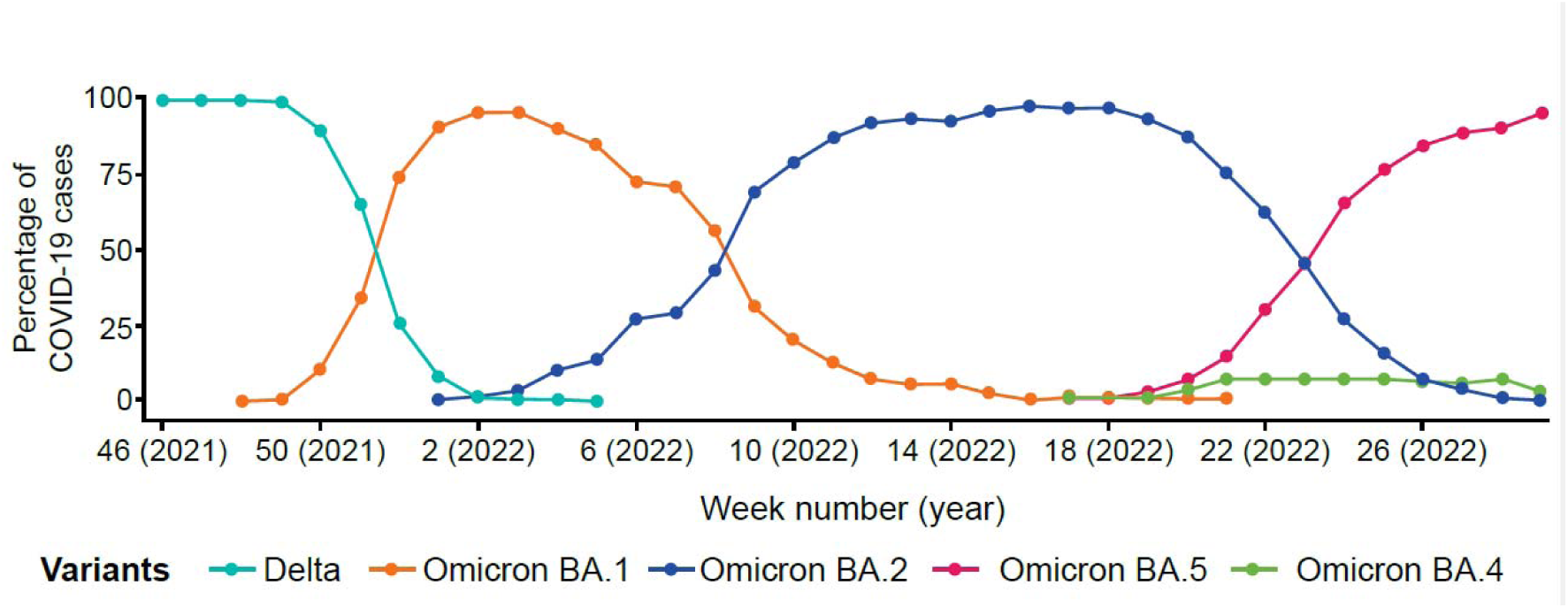
Proportion of SARS-CoV-2 variants over time in Belgium [28]. SARS-CoV-2=severe acute respiratory syndrome coronavirus 2

In addition, the length of stay (defined as number of hospitalization days), time to hospitalization after the index date, number and proportion of patients admitted to intensive care, and the number and proportion of patients receiving non-invasive or mechanical ventilation during all-cause or COVID-19-related hospitalization were captured during the acute period.

In an exploratory analysis, all-cause and COVID-19-related new hospitalizations and all-cause deaths reported during the post-acute period, defined as the period between Day 30 and Day 90 after the index date, were assessed. Data for the exploratory analysis could only be collected for patients identified on or before 1 June 2022 to allow time for Day 30–90 follow-up to be completed before the end of the study period (31 August 2022).

### Data analysis

Descriptive analyses were performed. For continuous variables, the mean, standard deviation (SD), median and interquartile range (IQR) were calculated. Proportions were calculated using the total number of eligible patients treated with sotrovimab as the denominator, unless otherwise specified. The prevalence of hospitalization or death during the respective SARS-CoV-2 variant predominance periods was calculated as the number of patients who experienced that event during the variant predominance period divided by the total number of sotrovimab-treated patients during that period multiplied by 100.

For post-acute 90-day outcomes, the proportions of hospitalizations (all-cause and COVID-19-related) and deaths (all-cause) were calculated among the number of patients treated with sotrovimab who were still alive at the beginning of the post-acute period and had completed the 90-day follow-up.

## Results

### Patient identification and characteristics

Following application of the inclusion and exclusion criteria, 634 patients were included in the study (**Table 1**). A total of 598 patients identified across the study centres had received sotrovimab while already hospitalized and were excluded from the study. No patients identified had received remdesivir as an outpatient during the acute COVID-19 infection period.

**Table 1.**
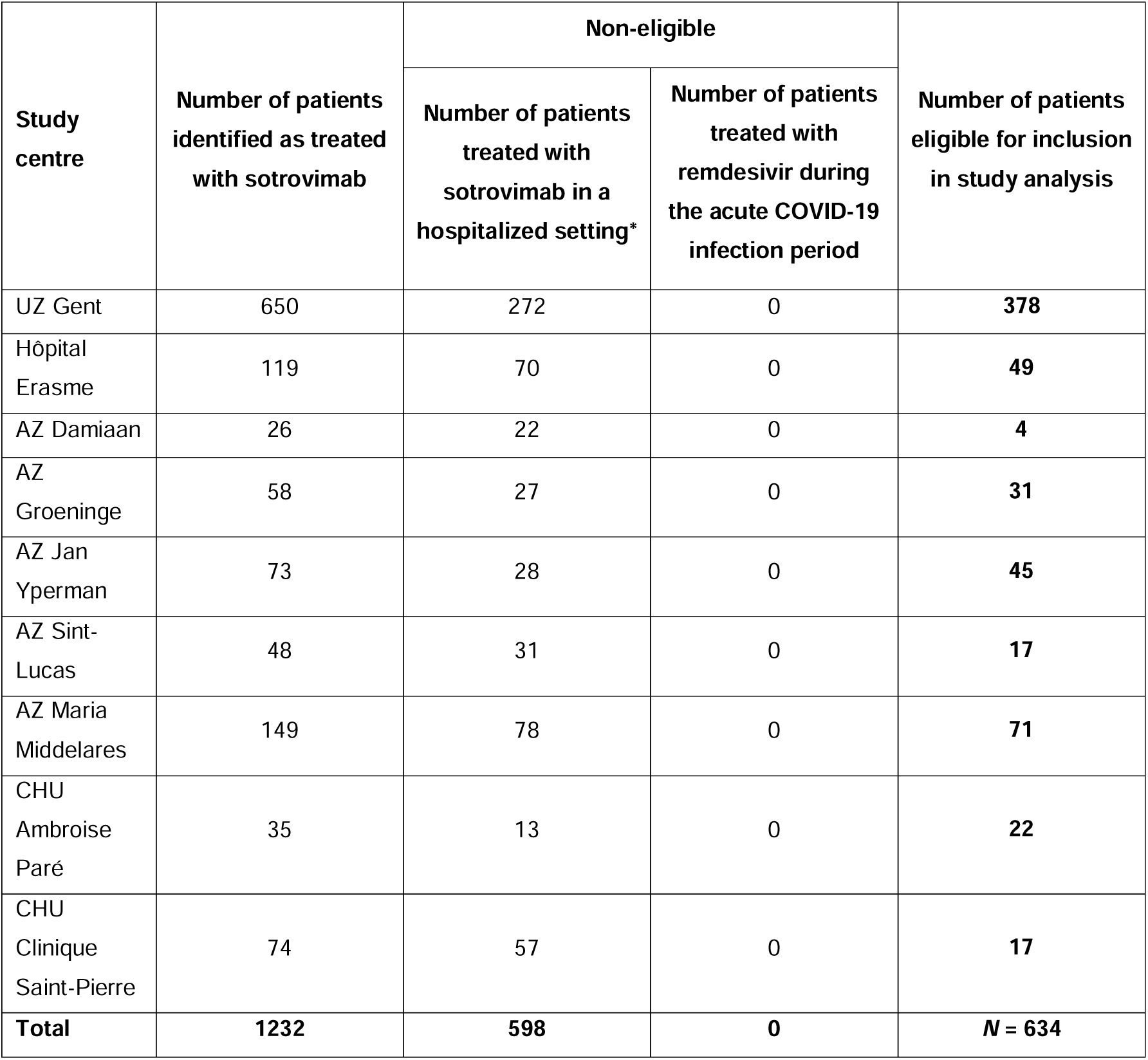
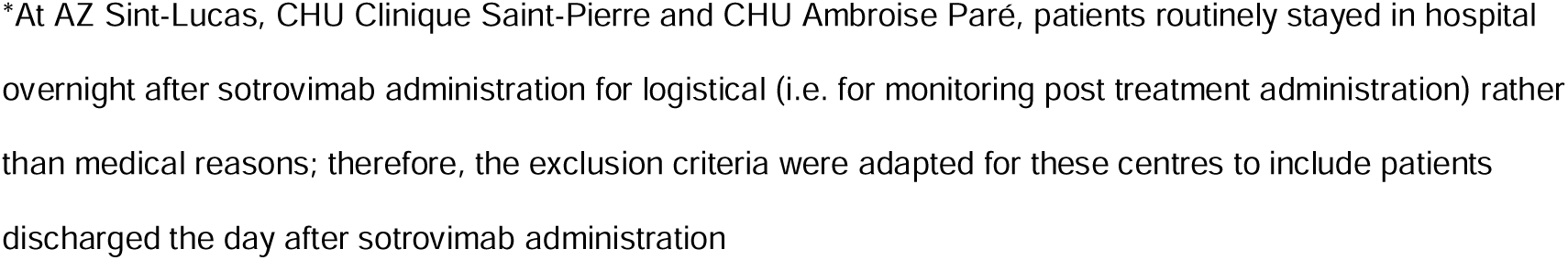
Number of patients excluded and included per study centre (1 November 2021*–*2 August 2022)

**Table 2** presents the baseline demographic and clinical characteristics among sotrovimab-treated patients included in the analysis. The patients were predominantly aged <65 years (63.4%, *n* = 402/634), with more than one-third (38.6%, *n* = 245/634) of patients aged 18–54 years. Half of the patients (50.3%, *n* = 319/634) were male, and 67.7% (*n* = 429/634) were immunocompromised and thus considered at high risk of developing COVID- 19 complications. Most of the immunocompromised patients had a malignancy and were undergoing treatment with antineoplastic agents, endocrine therapies or immunostimulants (36.9%, *n* = 234/634), or had primary immune deficiency (29.7%, *n* = 188/634) or a haematological malignancy (25.4%, *n* = 161/634). With respect to the other risk factors and comorbidities of interest (excluding age ≥65 years), cardiovascular disease (17.7%, *n* = 112/634), diabetes (12.3%, *n* = 78/634) and CKD (10.6%, *n* = 67/634) were the most common.

**Table 2.** Patient characteristics.

| Characteristic | Sotrovimab<br>($N = 634$) |
| --- | --- |
| **Age group, years, $n$ (%)** |  |
| 12–17 | 23 (3.6) |
| 18–54 | 245 (38.6) |
| 55–64 | 134 (21.1) |
| 65–74 | 140 (22.1) |
| $\geq 75$ | 92 (14.5) |
| **Male sex, $n$ (%)** | 319 (50.3) |
| **Immunocompromised patients at high risk of developing COVID-19 complications, $n$ (%)** |  |
| $\geq 1$ high-risk criterion | 429 (67.7) |
| HIV/AIDS | 4 (0.6) |
| Solid tumour | 85 (13.4) |
| Cancer undergoing treatment: radiotherapy | 0 |
| Cancer undergoing treatment: radio- or chemotherapy | 8 (1.3) |
| Cancer undergoing treatment: drugs | 234 (36.9) |
| Haematological malignancy | 161 (25.4) |
| Solid organ or haematopoietic stem cell transplantation | 121 (19.1) |
| Primary immune deficiency | 188 (29.7) |
| Immunosuppressive drugs | 120 (18.9) |
| Sickle cell anaemia | 3 (0.5) |
| Major thalassaemia | 0 |
| Asplenia | 0 |
| **Other risk factors/comorbidities of interest, n (%)** |  |
| ≥1 other comorbidity of interest | 364 (57.4) |
| Age ≥65 years | 232 (36.6) |
| Obesity with BMI ≥30 kg/m<sup>2</sup> | 42 (6.6) |
| Type 1 or type 2 diabetes mellitus | 78 (12.3) |
| Chronic lung disease, including asthma | 36 (5.7) |
| Cardiovascular disease, including uncontrolled hypertension | 112 (17.7) |
| Chronic kidney disease (eGFR <30 ml/min), including haemodialysis | 67 (10.6) |
| Chronic liver disease (Child–Pugh B or C) | 30 (4.7) |
| Chronic neurological disease | 23 (3.6) |

AIDS=acquired immunodeficiency syndrome; BMI=body mass index; COVID-19=coronavirus disease 2019;
eGFR=estimated glomerular filtration rate; HIV=human immunodeficiency virus

### Overall analysis of acute period (29-day) clinical outcomes

#### All-cause clinical outcomes

Among the 634 sotrovimab-treated patients, 82 (12.9%) were hospitalized or died due to any cause within the 29-day acute period (**Table 3**). Seventy-nine (12.5%) patients were hospitalized for any cause and seven (1.1%) died due to any cause during the 29-day acute period. Of the 79 patients who had all-cause hospitalization events, 12 (15.2%) had more than one hospitalization event during the acute phase. Among hospitalized patients, the median (IQR) duration of the first all-cause hospitalization was 4.0 (2.0, 9.0) days. Median (IQR) time to first all-cause hospitalization from the index date was 14.0 (9.5, 22.0) days.

**Table 3.** Acute period (29-day) all-cause and COVID-19-related outcomes.

| Outcome | Patients treated with sotrovimab<br>($N = 634$) |
| --- | --- |
| All-cause hospitalization or death, $n$ (%) | 82 (12.9) |
| All-cause hospitalization, $n$ (%) | 79 (12.5) |
| All-cause hospitalization events, $n$ (%) | |
| 0 | 555 (87.5) |
| 1 | 67 (10.6) |
| 2 | 10 (1.6) |
| 3 | 2 (0.3) |
| All-cause deaths, $n$ (%) | 7 (1.1) |
| Number of all-cause hospitalization days |  |
| Mean (SD) | 8.3 (12.5) |
| Median (IQR) | 4.0 (7.0) |
| Time to hospitalization from index date in days |  |
| Mean (SD) | 14.9 (7.8) |
| Median (IQR) | 14.0 (12.5) |
| All-cause ICU admission, $n$ (%) | 5 (0.8) |
| Number of ICU days |  |
| Mean (SD) | 7.2 (3.8) |
| Median (IQR) | 6.0 (5.0) |
| All-cause non-invasive ventilation, $n$ (%) | 1 (0.2) |
| All-cause mechanical ventilation, $n$ (%) | 2 (0.3) |
| COVID-19-related hospitalization, $n$ (%) | 16 (2.5)* |
| Number of COVID-19-related hospitalization days |  |
| Mean (SD) | 14.0 (16.3) |
| Median (IQR) | 10.0 (7.5) |
| Time to COVID-19-related hospitalization from index date in days | 11.4 (8.9) |
| Mean (SD)<br>Median (IQR) | 10.5 (18.5) |
| **COVID-19-related ICU admission, *n* (%)** | 3 (0.5) |
| **Number of COVID-19-related ICU days**<br>Mean (SD)<br>Median (IQR) | 5.7 (2.9)<br>4.0 (2.5) |
| **COVID-19-related non-invasive ventilation, *n* (%)** | 0 |
| **COVID-19-related mechanical ventilation, *n* (%)** | 1 (0.2) |

\*Fifteen patients (2.4%) had a first COVID-19-related hospitalization during the 29-day acute period. One patient was hospitalized for the first time during the 29-day acute period for reasons unrelated to COVID-19, but a subsequent hospitalization during the same period was COVID-related. COVID-19=coronavirus disease 2019, ICU=intensive care unit, IQR=interquartile range, SD=standard deviation

Five (6.3%) of the patients with all-cause hospitalizations were admitted to the intensive care unit (ICU), with a median (IQR) stay of 6.0 (4.0, 9.0) ICU days. Two patients required mechanical ventilation and one patient required non-invasive ventilation.

#### COVID-19-related clinical outcomes

COVID-19-related hospitalizations during the 29-day acute period were experienced by 2.5% (*n* = 16/634) of patients (**Table 3**). The median (IQR) duration of stay was 10.0 (6.8, 14.2) days. Median (IQR) time to hospitalization from the index date was 10.5 (2.2, 20.8) days. Of patients with COVID-19-related hospitalizations, three (18.8%) were admitted to an ICU, with a median (IQR) stay of 4.0 (4.0, 6.5) ICU days. One patient required mechanical ventilation.

Of the 378 patients treated with sotrovimab at Ghent University Hospital, four (1.1%) died due to any cause, including one patient who died as a result of COVID-19 complications.

### Acute period (29-day) clinical outcomes during periods of Delta and Omicron predominance

The 29-day acute period all-cause and COVID-19-related outcomes by period of SARS-CoV-2 variant predominance are shown in **Table 4**. The largest number of sotrovimab-treated patients were observed during the BA.1 (34.4%, *n* = 218/634) and BA.2/BA.5 codominance (40.2%, *n* = 255/634) periods.

**Table 4.**
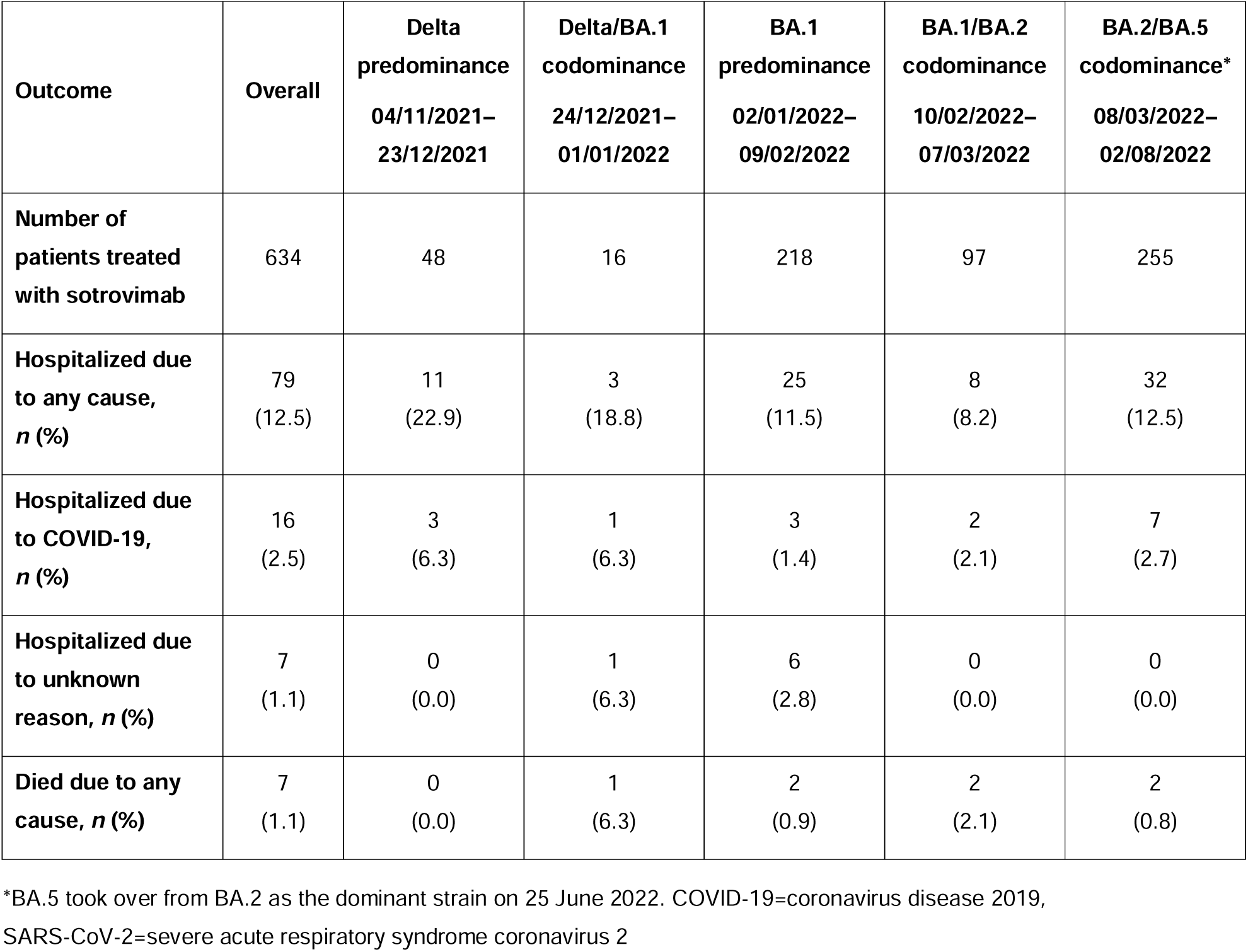
Acute period (29-day) all-cause and COVID-19-related outcomes by period of SARS-CoV-2 variant predominance.

| Outcome | Overall | Delta<br>predominance<br>04/11/2021–<br>23/12/2021 | Delta/BA.1<br>codominance<br>24/12/2021–<br>01/01/2022 | BA.1<br>predominance<br>02/01/2022–<br>09/02/2022 | BA.1/BA.2<br>codominance<br>10/02/2022–<br>07/03/2022 | BA.2/BA.5<br>codominance*<br>08/03/2022–<br>02/08/2022 |
| --- | --- | --- | --- | --- | --- | --- |
| Number of<br>patients treated<br>with sotrovimab | 634 | 48 | 16 | 218 | 97 | 255 |
| Hospitalized due<br>to any cause,<br>$n$ (%) | 79<br>(12.5) | 11<br>(22.9) | 3<br>(18.8) | 25<br>(11.5) | 8<br>(8.2) | 32<br>(12.5) |
| Hospitalized due<br>to COVID-19,<br>$n$ (%) | 16<br>(2.5) | 3<br>(6.3) | 1<br>(6.3) | 3<br>(1.4) | 2<br>(2.1) | 7<br>(2.7) |
| Hospitalized due<br>to unknown<br>reason, $n$ (%) | 7<br>(1.1) | 0<br>(0.0) | 1<br>(6.3) | 6<br>(2.8) | 0<br>(0.0) | 0<br>(0.0) |
| Died due to any<br>cause, $n$ (%) | 7<br>(1.1) | 0<br>(0.0) | 1<br>(6.3) | 2<br>(0.9) | 2<br>(2.1) | 2<br>(0.8) |

\*BA.5 took over from BA.2 as the dominant strain on 25 June 2022. COVID-19=coronavirus disease 2019,
SARS-CoV-2=severe acute respiratory syndrome coronavirus 2

During the Delta predominance period, the proportion of sotrovimab-treated patients hospitalized due to any cause was 22.9% (*n* = 11/48). During the Delta/BA.1 codominance, BA.1 predominance, BA.1/BA.2 codominance and BA.2/BA.5 codominance periods, all-cause hospitalizations were experienced by 18.8% (*n* = 3/16), 11.5% (*n* = 25/218), 8.2% (*n* = 8/97) and 12.5% (*n* = 32/255) of patients, respectively.

COVID-19-related hospitalizations were experienced by 6.3% (*n* = 3/48) of sotrovimab-treated patients during the period of Delta predominance, 6.3% (*n* = 1/16) in the Delta/BA.1 codominance period, 1.4% (*n* = 3/218) in the BA.1 predominance period, 2.1% (*n* = 2/97) in the BA.1/BA.2 codominance period and 2.7% (*n* = 7/255) in the BA.2/BA.5 codominance period (**Table 4**).

No sotrovimab-treated patients died during the Delta predominance period (**Table 4**). During the Delta/BA.1 codominance period, one sotrovimab-treated patient died (*n* = 1/16). Two patients died in each of the BA.1 predominance (*n* = 2/218), BA.1/BA.2 codominance (*n* = 2/97) and BA.2/BA.5 codominance (*n* = 2/255) periods.

### Exploratory analysis of post-acute period (30–90-day) clinical outcomes

In total, 625 patients had follow-up data available for the post-acute period, defined as 30– 90 days after the first sotrovimab administration (index date). Among these patients, 13.8% (*n* = 86/625) had a new hospitalization event (any cause), 1.6% (*n* = 10/625) had a COVID-19-related hospitalization and 1.1% (*n* = 7/625) died due to any cause during the post-acute period.

At Ghent University Hospital, 30–90-day follow-up data were available for 374 patients. Among these patients, five died (1.3%), with two deaths due to COVID-19-related reasons.

## Discussion

This single-arm, retrospective study describes the real-world clinical outcomes of patients with COVID-19 who received sotrovimab treatment across nine hospitals in Belgium between 1 November 2021 and 31 August 2022. Over two-thirds (67.7%) of the patients in this study had at least one immunocompromising condition, with over one-third of patients (36.9%) receiving chemotherapy for an underlying malignancy. Other commonly reported risk factors for progression to severe COVID-19 were age ≥65 years old, cardiovascular diseases (including uncontrolled hypertension), diabetes and CKD. Comparatively, national hospital surveillance, conducted by Sciensano, identified hypertension, cardiovascular disease and diabetes as the top three most frequently reported comorbidities among patients with COVID-19 hospitalized in Belgium over the course of the pandemic [29].

During this study, 12.9% of patients (*n* = 82/634) treated with sotrovimab were hospitalized or died due to any cause during the 29-day acute period. This rate is relatively high compared with rates reported in other retrospective or observational studies (all-cause hospitalization and/or mortality ranged from 2.1% to 2.7% during the BA.1 predominance period and from 1.7% to 2.0% during the BA.2 predominance period in a recent systematic literature review reporting on the real-world effectiveness of sotrovimab [30]), but this may be potentially explained by several observations. Firstly, high proportions of all-cause and COVID-19-related hospitalizations were mainly observed among sotrovimab-treated patients during the Delta predominance period (4 November to 23 December 2021) in Belgium.

These observations align with nationally reported records of high rates of COVID-19-related hospitalization (12,588 admissions) and deaths (1935) during the Delta predominance period [3, 31] and may reflect the higher disease severity, higher prevalence and more rigorous public health measures associated with the Delta variant relative to subsequent Omicron variants, as well as lower immunity within the population [32, 33]. Additionally, the present study period spanned four SARS-CoV-2 variants (Delta, BA.1, BA.2 and BA.5) compared with one to two variants (primarily BA.1 or BA.2) in other studies [34–36]. Furthermore, the current study had a relatively high proportion of patients with at least one immunocompromising condition (67.7%) compared with other studies [34, 35, 37, 38]. This higher proportion of high-risk patients, necessitating regular inpatient treatment for their underlying conditions, may explain the relatively higher rate of all-cause hospitalization or mortality observed in this current study compared with other real-world studies [30, 34, 35, 39].

In a recent Danish cohort study, 83% of 2933 sotrovimab-treated patients belonged to very-high-risk groups that included haematological malignancy, solid organ transplantation, and treatment with anti-CD20 for non-malignant haematological diseases within the previous year. Within 90 days of sotrovimab treatment (696 person-years), 30.2% of patients were hospitalized and 5.3% had died due to any cause. The highest prognostic factor was haematological malignancy, followed by age ≥65 years [40]. Comparatively, in our study, 68% of patients had at least one immunocompromising condition. A quarter (25.4%) of patients had a haematological malignancy and 36.6% were aged ≥65 years; our 30–90-day results show that 13.8% of patients had a new hospitalization event (any cause), and 1.1% died due to any cause. Similar to our findings, the Danish study found a higher proportion of hospitality and death associated with the Delta variant, but no major differences in these outcomes were observed between the Omicron BA.1 and BA.2 variants [40].

In our study population, the proportion of sotrovimab-treated patients experiencing COVID-19-related hospitalization was highest during the Delta predominance and Delta/BA.1 codominance (both 6.3%) periods. During the BA.1 predominance, BA.1/BA.2 codominance and BA.2/BA.5 codominance periods, COVID-19-related hospitalizations were consistently low (all ≤2.7%). The median duration of hospitalization in our study (10.0 days) was higher than the national reported figure for Belgium, of 2 days, during the Omicron predominance period [41]; however, the median duration of ICU stays (4.0 days) was the same as the national median duration reported for Belgium [41]. It should be noted that the number of patients admitted to an ICU was low (*n* = 3), which limits interpretation.

This study has several potential limitations that should be considered. Firstly, this descriptive, single-arm study did not include a comparator arm, such as patients with similar risk profiles who did not receive sotrovimab after SARS-CoV-2 infection. We were not able to derive a control group due to a lack of similarly high-risk patients who had not received sotrovimab treatment and because patients who did not receive sotrovimab and were not hospitalized were not accounted for in the data used in this study. As a result, it is difficult to infer an association between sotrovimab treatment and clinical outcomes from these data alone. Due to the retrospective, observational study design, variables such as vaccination status and disease severity at enrolment could not be extracted, as these were not structurally reported and standardized in all hospitals. However, vaccination rates in Belgium had reached 73% by the start of this study [42] (and were presumably higher in high-risk groups similar to this study population). Furthermore, one might assume a high vaccination uptake in this population, with patients having received regular follow-up at their treating hospital and being eligible for early treatment with mAbs. Patients who received sotrovimab as an inpatient were excluded from the study, with the exception of patients who were discharged the day after sotrovimab administration. This comprised a large proportion of the total number of patients identified (48.5% [*n* = 598/1232], potentially inducing selection bias in the analyses and limiting the generalizability of the findings. In addition, patients who received sotrovimab in one of the selected hospitals but were later admitted to a different hospital where they progressed to severe disease may not have been captured in our study given its retrospective nature. However, as sotrovimab is expected to be primarily used in patients at risk of progressing to severe outcomes, such as those who are immunocompromised, these patients were likely associated with a reference hospital given their need for frequent medical care; therefore, missing data for clinical outcomes were likely to be low. Similarly, data on the uptake of immunosuppressant drugs and diagnosis of comorbidities were only extractable for patients who were treated or diagnosed in the same hospital where they received their sotrovimab treatment. Information on drugs received in another hospital or prescribed in primary care were not available. Therefore, the number of patients who were at high risk of developing severe COVID-19 disease may have been underestimated. Lastly, as variant sequencing was not routinely undertaken, the specific SARS-CoV-2 variant with which patients were infected could not be determined. Instead, an ecological design was proposed that used the periods of Delta, BA.1, BA.2 and BA.5 subvariant predominance and codominance as a proxy for the infecting variant. National genomic surveillance data were used to stratify the study periods into circulating variant predominance periods [28].

## Conclusions

This observational study demonstrated consistently low rates of COVID-19-related hospitalizations and all-cause deaths in sotrovimab-treated patients during the Omicron subvariant predominance periods in Belgium, despite the reported decrease in *in vitro* neutralization activity of sotrovimab against BA.2 and BA.5, and over two-thirds of the study population being immunocompromised. Comparative effectiveness studies are warranted to confirm the effectiveness of sotrovimab in highly immunocompromised patients with COVID-19.

## Data Availability

The datasets used and/or analysed during the current study are not available due to data privacy.

## List of abbreviations

ATC: Anatomical Therapeutic Chemical
CKD: chronic kidney disease
COVID-19: coronavirus disease 2019
EMA: European Medicines Agency
ICD-10: International Classification of Disease version 10
ICU: intensive care unit
IQR: interquartile range
mAb: monoclonal antibody
SARS-CoV-2: severe acute respiratory syndrome coronavirus 2
SD: standard deviation

## Acknowledgements

The authors wish to thank the Ghent University Hospital Data Science Institute for its support and role in this study.

Editorial support (in the form of writing assistance, including preparation of the draft manuscript under the direction and guidance of the authors, collating and incorporating authors’ comments for each draft, assembling tables, grammatical editing and referencing) was provided by Kathryn Wardle and Tony Reardon of Apollo, OPEN Health Communications, in accordance with Good Publication Practice guidelines (www.ismpp.org/gpp-2022). The support was funded by GSK and Vir Biotechnology, Inc.

## Declarations

### Authors’ contributions

Conception: MD, VP, DCG, EJL, JH, LL

Study design: MD, VP, DCG, EJL, JH, LL, TH, KC, EVB

Execution: MD, VP, DCG, EJL, JH, LL, TH, KC, EVB, MS

Acquisition of data: TH, KC, EVB

Analysis and interpretation: MD, VP, DCG, EJL, JH, LL, TH, KC, EVB, MS

All authors took part in drafting, revising or critically reviewing the manuscript; gave final approval of the version to be published; have agreed on the journal to which the article has been submitted; and agree to be accountable for all aspects of the work.

### Availability of data and materials

The datasets used and/or analysed during the current study are not available due to data privacy.

### Ethics approval and consent to participate

This study complies with all applicable laws regarding subject privacy. No direct subject contact or primary collection of individual human subject data occurred. Study results were in tabular form and were aggregate analyses that omit subject identification; therefore, informed consent was not required. Ethics committee approval was obtained from each of the participating centres (CHU Saint-Pierre - Comité local d’éthique hospitalier [n° 007]; CHU Ambroise Paré - Comité d’éthique hospitalier; AZ Damiaan - Ethisch comité [n° 057]; AZ Sint Lucas Gent - Commissie medische ethiek; Hôpital Erasme ULB - Comité d’éthique hospitalo-facultaire; UZ Gent - Commissie voor medische ethiek), with the exception of AZ Maria Middelares, AZ Groeninge and Jan Yperman. For AZ Maria Middelares, formal ethics committee approval is not required for retrospective database studies; however, the study was approved by a data protection board that oversees information security, ethical and financial aspects in the context of secondary reuse of health data. For AZ Groeninge and Jan Yperman, formal ethics approval is not required for retrospective studies; however, a data access committee at each centre (with medical legal and ethics committee representation) approved the study.

### Consent for publication

Not applicable.

## Additional files

Additional file 1 (PDF) - Supplementary Appendix, containing ATC and ICD-10 codes used in this study, and all-cause and COVID-19-related outcomes by study month

