## Additional File for "Characteristics and outcomes of patients treated with sotrovimab to prevent progression to severe COVID-19 in Belgium"

### Supplementary Appendix

**Table S1** List of immunosuppressant and cancer treatment drugs

| Drug name | ATC code |
|---|---|
| **Immunosuppressant drugs** | |
| 6-mercaptopurine | L01BB02 |
| Abatacept | L04AA24 |
| Adalimumab | L04AB04 |
| Alemtuzumab | L04AA34 |
| Anakinra | L04AC03 |
| Apremilast | L04AA32 |
| Azathioprine | L04AX01 |
| Baricitinib | L04AA37 |
| Basiliximab | L04AC02 |
| Belatacept | L04AA28 |
| Belimumab | L04AA26 |
| Bendamustine | L01AA09 |
| Brentuximab | L01FX05 |
| Busulfan | L01AB01 |
| Canakinumab | L04AC08 |
| Certolizumab | L04AB05 |
| Chlorambucil | L01AA02 |
| Cyclophosphamide | L01AA01 |
| Cyclosporine | L04AD01 |
| Dacarbazine | L01AX04 |
| Daclizumab | L04AC01 |
| Dimethyl fumarate | L04AX07, N07XX09 |
| Eculizumab | L04AA25 |
| Elotuzumab | L01FX08 |
| Estramustine | L01XX11 |
| Etanercept | L04AB01 |
| Everolimus | L04AA18, L01EG02, L01EG01 |
| Fingolimod | L04AA27 |
| Fotemustine | L01AD05 |
| Glatiramer acetate | L03AX13 |
| Golimumab | L04AB06 |
| Ifosfamide | L01AA06 |

| | |
|---|---|
| Infliximab | L04AB02 |
| Ipilimumab | L01FX04 |
| Ixekizumab | L04AC13 |
| Leflunomide | L04AA13 |
| Melphalan | L01AA03 |
| Methotrexate | L01BA01, L04AX03 |
| Mitoxantrone | L01DB07 |
| Mycophenolate mofetil | L04AA06 |
| Natalizumab | L04AA23 |
| Obinutuzumab | L01FA03 |
| Prednisone (≥20 mg for ≥14 days) | S01BA04, H02AB06, A07EA01, C05AA04, R01AD02, S01BA03 |
| Rituximab | L01FA01 |
| Secukinumab | L04AC10 |
| Siltuximab | L04AC11 |
| Sirolimus | L04AA10, S01XA23 |
| Tacrolimus | L04AD02 |
| Temozolomide | L01AX03 |
| Teriflunomide | L04AA31 |
| Thiotepa | L01AC01 |
| Tocilizumab | L04AC07 |
| Tofacitinib | L04AA29 |
| Ustekinumab | L04AC05 |
| Vedolizumab | L04AA33 |
| **Cancer treatment: radiotherapy drugs** | |
| Radium dichloride | V10XX03 |
| **Cancer treatment: drugs** | |
| Antineoplastic agents and immunomodulating agents | L01 |
| Endocrine therapy | L02 |
| Immunostimulants | L03 |

ATC=Anatomical Therapeutic Chemical.

**Table S2** List of ICD-10 codes to identify immunocompromised patients and other comorbidities

| ICD-10 codes | Description |
|---|---|
| **HIV** | |
| B20 | Human immunodeficiency virus (HIV) disease |
| B97.35 | Human immunodeficiency virus, type 2 (HIV-2) |
| **Primary immune deficiency (disorders involving the immune mechanism)** | |
| D50–D89 | Diseases of the blood and blood-forming organs and certain disorders involving the immune mechanism |
| K50 | Crohn's disease (regional enteritis) |
| K51.90 | Ulcerative enterocolitis |
| K52.9 | Other and unspecified non-infectious gastroenteritis and colitis |
| L40 | Psoriasis |
| M06.4 | Rheumatoid arthritis and other inflammatory polyarthropathies |
| M32.9 | Systemic lupus erythematosus |
| Q89.0, Q89.01 | Asplenia (congenital) |
| **Transplant** | |
| T86 | Complications of transplanted organs and tissue |
| Z94 | Transplanted organ and tissue status |
| **Solid tumours** | |
| C00–C14 | Malignant neoplasms of lip, oral cavity and pharynx |
| C15–C26 | Malignant neoplasms of digestive organs |
| C30–C39 | Malignant neoplasms of respiratory and intrathoracic organs |
| C40–C41 | Malignant neoplasms of bone and articular cartilage |
| C43–C44 | Melanoma and other malignant neoplasms of skin |
| C45–C49 | Malignant neoplasms of mesothelial and soft tissue |
| C50 | Malignant neoplasms of breast |
| C51–C58 | Malignant neoplasms of female genital organs |
| C60–C63 | Malignant neoplasms of male genital organs |
| C64–C68 | Malignant neoplasms of urinary tract |
| C69–C72 | Malignant neoplasms of eye, brain and other parts of central nervous system |
| C73–C75 | Malignant neoplasms of thyroid and other endocrine glands |
| C76–C80 | Malignant neoplasms of ill-defined, other secondary and unspecified sites |
| C7A | Malignant neuroendocrine tumours |
| C7B | Secondary neuroendocrine tumours |
| D00–D09 | *In situ* neoplasms |

| | |
|---|---|
| D49 | Neoplasms of unspecified behaviour |
| D37–D44 | Neoplasms of uncertain behaviour |
| D48 | Neoplasm of uncertain behaviour of other and unspecified sites |
| **Haematological malignancies** | |
| C81–C96 | Malignant neoplasms of lymphoid, haematopoietic and related tissue |
| D45–D47 | Polycythaemia vera, myelodysplastic syndromes and other neoplasms of uncertain behaviour of lymphoid, haematopoietic and related tissue |
| **Sickle cell anaemia** | |
| D57.0 | Hb-SS disease with crisis |
| D57.1 | Sickle cell disease without crisis |
| D57.2 | Sickle cell/Hb-C disease |
| **Major thalassaemia** | |
| D56.0 | Alpha thalassaemia |
| D56.1 | Beta thalassaemia |
| **Chronic kidney disease** | |
| N17 | Acute kidney failure |
| N18.4 | CKD Stage 4 |
| N18.5 | CKD Stage 5 |
| N18.6 | ESRD |
| Z99.2 | Dependence on renal dialysis |
| Z49 | Encounter for other dialysis |
| Z49.01 | Fitting and adjustment of extracorporeal dialysis catheter |
| Z49.02 | Peritoneal dialysis |
| Z49.31 | Encounter for adequacy testing for haemodialysis |
| Z49.32 | Encounter for adequacy testing for peritoneal haemodialysis |
| **Chronic liver disease** | |
| I85.10 | Oesophageal varices in diseases classified elsewhere, without mention of bleeding |
| K72.00 | Acute and subacute necrosis of liver |
| K72.10 | Other sequelae of chronic liver disease |
| K73 | Chronic hepatitis, not elsewhere classified |
| K72.91 | Hepatic encephalopathy |
| K73.9 | Hepatitis, unspecified |
| K74 | Fibrosis and cirrhosis of liver |
| K76.6 | Portal hypertension |
| K76.7 | Hepatorenal syndrome |
| K76.81 | Hepatopulmonary syndrome |

| | |
|---|---|
| K76.89 | Liver disorders |
| K76.9 | Unspecified disorder of liver |
| K76.1 | Chronic passive congestion of liver |
| R18.8 | Other ascites |
| **Type 1 or type 2 diabetes** | |
| E08 | Diabetes mellitus due to underlying condition |
| E09 | Drug or chemical induced diabetes mellitus |
| E10 | Type 1 diabetes mellitus |
| E11 | Type 2 diabetes mellitus |
| E13 | Other specified diabetes mellitus |
| **Obesity** | |
| E66.0 | Obesity due to excess calories |
| E66.1 | Drug-induced obesity |
| E66.2 | Morbid (severe) obesity with alveolar hypoventilation |
| E66.8 | Other obesity |
| E66.9 | Obesity, unspecified |
| **Chronic cardiovascular disease** | |
| I20–I25 | Ischaemic heart diseases |
| I27.0 | Primary pulmonary hypertension |
| I27.2 | Other secondary pulmonary hypertension |
| I27.81 | Cor pulmonale (chronic) |
| I27.9 | Pulmonary heart disease, unspecified |
| I5A | Non-ischaemic myocardial injury (nontraumatic) |
| I30–I52 | Other forms of heart disease |
| Q20–Q28 | Congenital malformations of the circulatory system |
| **Chronic lung disease** | |
| E84.9 | Cystic fibrosis, unspecified |
| E84.11 | Meconium ileus in cystic fibrosis |
| I26 | Pulmonary embolism |
| E84.0 | Cystic fibrosis with pulmonary manifestations |
| I27.82 | Chronic pulmonary embolism |
| J44.9 | Chronic obstructive pulmonary disease |
| J45.901 | Unspecified asthma with (acute) exacerbation |
| J45.902 | Unspecified asthma with status asthmaticus |
| J84.10 | Pulmonary fibrosis, unspecified |
| J84.89 | Other specified interstitial pulmonary diseases |
| J98.4 | Other disorders of lung |
| **Chronic neurological disease** | |
| A81.01 | Variant Creutzfeldt–Jakob disease |

| | |
|---|---|
| F01.50 | Vascular dementia without behavioural disturbance |
| F07.0 | Personality change due to known physiological condition |
| G10 | Huntington's disease |
| G20 | Parkinson's disease |
| G21 | Secondary parkinsonism |
| G23 | Other degenerative diseases of basal ganglia |
| G24 | Dystonia |
| G25 | Other extrapyramidal and movement disorders |
| G26 | Extrapyramidal and movement disorders in diseases classified elsewhere |
| G30 | Alzheimer's disease |
| G31 | Other degenerative diseases of nervous system, not elsewhere classified |
| G32 | Other degenerative disorders of nervous system in diseases classified elsewhere |
| I60 | Nontraumatic subarachnoid haemorrhage |
| I61 | Nontraumatic intracerebral haemorrhage |
| I62 | Other and unspecified nontraumatic intracranial haemorrhage |
| I63 | Cerebral infarction |
| I65 | Occlusion and stenosis of precerebral arteries, not resulting in cerebral infarction |
| I66 | Occlusion and stenosis of cerebral arteries, not resulting in cerebral infarction |
| I67 | Other cerebrovascular diseases |
| I68 | Cerebrovascular disorders in diseases classified elsewhere |
| I69 | Sequelae of cerebrovascular disease |
| R47.01 | Aphasia |
| R40.20 | Unspecified coma |
| **Cancer treatment: radiotherapy or chemotherapy** | |
| Z51.0 | Encounter for antineoplastic radiation therapy |
| Z51.1 | Encounter for antineoplastic chemotherapy and immunotherapy |
| **COVID-19-related hospitalizations** | |
| B34.2 | Coronavirus infection, unspecified |
| U07.1 | COVID-19 |
| B97.2 | Coronavirus as the cause of diseases classified elsewhere |

CKD=chronic kidney disease, COVID-19=coronavirus disease 2019, ESRD=end-stage renal disease, HIV=human immunodeficiency virus, ICD-10=International Classification of Disease version 10

**Table S3** List of ICD-10 procedure codes

| ICD-10 codes | Description |
|---|---|
| **Invasive mechanical ventilation** | |
| 0BH13EZ | Insertion of endotracheal airway into trachea, percutaneous approach |
| 0BH17EZ | Insertion of endotracheal airway into trachea, via natural or artificial opening |
| 0BH18EZ | Insertion of endotracheal airway into trachea, via natural or artificial opening |
| 0B110F4 | Bypass trachea to cutaneous with tracheostomy device, open approach |
| 0B110Z4 | Bypass trachea to cutaneous, open approach |
| 0B113F4 | Bypass trachea to cutaneous with tracheostomy device, percutaneous approach |
| 0B113Z4 | Bypass trachea to cutaneous, percutaneous approach |
| 0B114F4 | Bypass trachea to cutaneous with tracheostomy device, percutaneous endoscopic approach |
| 0B114Z4 | Bypass trachea to cutaneous, percutaneous endoscopic approach |
| 5A1.935Z | Extracorporeal or systemic assistance and performance, physiological systems, performance, respiratory, less than 24 consecutive hours, ventilation, no qualifier |
| 5A1.945Z | Extracorporeal or systemic assistance and performance, physiological systems, performance, respiratory, 24–96 consecutive hours, ventilation, no qualifier |
| 5A1.955Z | Extracorporeal or systemic assistance and performance, physiological systems, performance, respiratory, greater than 96 consecutive hours, ventilation, no qualifier |
| **Non-invasive ventilation** | |
| 5A0.9357 | Assistance with respiratory ventilation, less than 24 consecutive hours, continuous positive airway pressure |
| 5A0.9358 | Assistance with respiratory ventilation, less than 24 consecutive hours, intermittent positive airway pressure |
| 5A0.9359 | Assistance with respiratory ventilation, less than 24 consecutive hours, continuous negative airway pressure |
| 5A0.935B | Assistance with respiratory ventilation, less than 24 consecutive hours, intermittent negative airway pressure |
| 5A0.935Z | Assistance with respiratory ventilation, less than 24 consecutive hours |

| | |
|---|---|
| 5A0.9457 | Assistance with respiratory ventilation, 24–96 consecutive hours, continuous positive airway pressure |
| 5A0.9458 | Assistance with respiratory ventilation, 24–96 consecutive hours, intermittent positive airway pressure |
| 5A0.9459 | Assistance with respiratory ventilation, 24–96 consecutive hours, continuous negative airway pressure |
| 5A0.945B | Assistance with respiratory ventilation, 24–96 consecutive hours, intermittent negative airway pressure |
| 5A0.945Z | Assistance with respiratory ventilation, 24–96 consecutive hours |
| 5A0.9557 | Assistance with respiratory ventilation, greater than 96 consecutive hours, continuous positive airway pressure |
| 5A0.9558 | Assistance with respiratory ventilation, greater than 96 consecutive hours, intermittent positive airway pressure |
| 5A0.9559 | Assistance with respiratory ventilation, greater than 96 consecutive hours, continuous negative airway pressure |
| 5A0.955B | Assistance with respiratory ventilation, greater than 96 consecutive hours, intermittent negative airway pressure |
| 5A0.955Z | Assistance with respiratory ventilation, greater than 96 consecutive hours |
| 5A1.9054 | Respiratory ventilation, dingle, nonmechanical |
| Z99.11 | Dependence on respirator (ventilator) status |

ICD-10=International Classification of Disease version 10

**Table S4** Acute period (29-day) all-cause and COVID-19-related outcomes by study month

| **Outcome** | **Study month** | | | | | | | | | |
|---|---|---|---|---|---|---|---|---|---|---|
| | **Overall** | **Nov 2021** | **Dec 2021** | **Jan 2022** | **Feb 2022** | **Mar 2022** | **Apr 2022** | **May 2022** | **Jun 2022** | **Jul 2022** |
| **Number of patients treated with sotrovimab** | 634 | 20 | 44 | 178 | 112 | 148 | 127 | 1 | 2 | 2 |
| **Hospitalized due to any cause, *n* (%)** | 79 (12.5) | 4 (20.0) | 10 (22.7) | 20 (11.2) | 9 (8.0) | 22 (14.9) | 14 (11.0) | 0 (0.0) | 0 (0.0) | 0 (0.0) |
| **Hospitalized due to COVID-19, *n* (%)** | 16 (2.5) | 0 (0.0) | 4 (9.1) | 2 (1.1) | 3 (2.7) | 4 (2.7) | 3 (2.4) | 0 (0.0) | 0 (0.0) | 0 (0.0) |
| **Hospitalized due to unknown reason, *n* (%)** | 7 (1.1) | 0 (0.0) | 1 (2.3) | 5 (2.8) | 1 (0.9) | 0 (0.0) | 0 (0.0) | 0 (0.0) | 0 (0.0) | 0 (0.0) |
| **Died due to any cause, *n* (%)** | 7 (1.1) | 0 (0.0) | 1 (2.3) | 2 (1.1) | 2 (1.8) | 2 (1.4) | 0 (0.0) | 0 (0.0) | 0 (0.0) | 0 (0.0) |

COVID-19=coronavirus disease 2019
